# Understanding genetic risk factors for common side effects of antidepressant medications

**DOI:** 10.1101/2021.07.12.21260397

**Authors:** Adrian I. Campos, Aoibhe Mulcahy, Jackson G. Thorp, Naomi R. Wray, Enda M. Byrne, Penelope A. Lind, Sarah E. Medland, Nicholas G. Martin, Ian B. Hickie, Miguel E. Rentería

## Abstract

Major depression is one of the most disabling health conditions internationally. In recent years, new generation antidepressant medicines have become very widely prescribed. While these medicines are efficacious, side effects are common and frequently result in discontinuation of treatment. Compared with specific pharmacological properties of the different medications, the relevance of individual vulnerability is understudied. We used data from the Australian Genetics of Depression Study to gain insights into the aetiology and genetic risk factors to antidepressant side effects. As expected, the most commonly reported longer-term side effects were reduced sexual function and weight gain. Importantly, participants reporting a specific side effect for one antidepressant were more likely to report the same side effect for other antidepressants, suggesting the presence of shared individual or pharmacological factors. Depression Polygenic Risk Scores (PRS) were associated with side effects that overlapped with depressive symptoms, including suicidality and anxiety. Body Mass Index PRS were strongly associated with weight gain from all medications. PRS for headaches were associated with headaches from sertraline. Insomnia PRS showed some evidence of predicting insomnia from amitriptyline and escitalopram. Our results suggest a set of common factors underlying the risk for antidepressant side effects. These factors seem to be, at least in part, explained by genetic liability related to depression severity and the nature of the side effect. Future studies on the genetic aetiology of side effects will enable insights into their underlying mechanisms and the possibility of risk stratification and prophylaxis strategies.

## Introduction

The World Health Organisation predicts that depression will become the leading cause of disability globally by 2030 (1). The symptomatology, longitudinal course, response to treatment and functional impact of depressive disorders are highly variable. Antidepressant medicines are widely prescribed across the spectrum of depression severity and subtypes, alone or in combination with psychological therapies.

Effective pharmacotherapies for depression were first developed in the 1960s, following the identification of antipsychotic therapies. A clear focus was on the regulation of brain monoamine systems (dopamine, serotonin and noradrenaline). These agents, including tricyclic antidepressants (TCAs) and Monoamine Oxidase Inhibitors (MAOIs), were limited in the extent of their use by considerable side effect burdens and potential toxicity. From the 1980s onwards, further pharmacological developments have been dominated by the establishment of second-generation antidepressant classes, including selective serotonin reuptake inhibitors (SSRIs) and serotonin-norepinephrine reuptake inhibitors (SNRIs) (2). Although second-generation antidepressants have been shown to alleviate depression (3), treatment response is heterogeneous, and new side effect profiles have emerged (gastrointestinal, weight gain, sexual dysfunction). The degree of individual variation in the incidence and severity of these difficulties is high.

Treatment failure is commonly caused by discontinuation of antidepressants from adverse side effects. Over half of individuals have been recorded to cease medication within the first six months of initial prescription (4). Previously reported antidepressant adverse effects include sexual dysfunction (5–7), weight changes (8–11), insomnia (12–15), and suicidality (16–18). However, these ‘side effects’ may also reflect ongoing symptoms of the depressive illness. For example, anhedonia is a cardinal symptom of major depressive disorder (MDD) which could explain lower levels of sexual interest and arousal leading to sexual function impairments (19,20). Weight changes, sleep disturbances, and suicidality are also symptoms of depression (21) and its various phenotypic subtypes. Finally, other comorbid mental health disorders may amplify or trigger suicidal behaviours (22). Whether these side effects stem from adverse reactions to antidepressants or whether they are extensions or exacerbations of characteristics of an individual’s depression or a consequence of comorbidity with another disorder remains unclear.

Variability in medication response and tolerability may be inherited. For instance, genetic variation leading to changes in the function of antidepressant metabolising enzymes are believed to underlie side effects due to drug overexposure (23). Five to seven percent of Caucasian individuals are estimated to be poor CYP2D6 metabolisers (24), one of the major metabolising enzymes of fluoxetine, paroxetine and fluvoxamine. Furthermore, variants in genes such as *CYP2C19* and *CYP3A4* have been linked to citalopram (25) and sertraline (26) differential metabolism and clearance. Drug metabolising enzymes are biologically-relevant hypotheses for understanding adverse side effects. Nonetheless, genetic variants within these enzymes have failed to reach significance in recent genome-wide association studies (GWAS) on treatment resistance (27) and response (28), suggesting that treatment outcomes might be more complex than previously thought. It is likely that genetic factors underlying antidepressant side effects are a product of drug-specific factors such as variation within drug-metabolizing enzymes, as well as common (or non drug-specific) factors, the nature of which remains elusive.

In general, the aetiology of antidepressant adverse side effects remains largely understudied. Thus, we aim to bridge this research gap by leveraging data from the Australian Genetics of Depression Study (AGDS) to gain insights into the prevalence, aetiology and genetic underpinnings of adverse side effects associated with antidepressant use. We investigate the prevalence and demographic risk factors for 23 side effects across ten commonly prescribed antidepressants. We test for SSRI or SNRI specificity and provide evidence for a co-occurring relationship between adverse side effects across different antidepressant medications. That is, participants who took two or more antidepressants were more likely to report the same side effects regardless of the antidepressant used. This co-occurrence would suggest a set of common risk factors underlie these side effects. Here, we use polygenic risk scores (PRS) to study the genetic aetiology of specific antidepressant adverse side effects to understand the nature of these common risk factors. PRS are an estimate of an individual’s genetic risk for a certain trait. They are calculated based on GWAS results whereby genetic variants are linked to a trait of interest through an effect size (i.e., the increased risk per copy of the genetic variant allele). PRS are calculated in an independent sample by performing a sum of risk variants weighted by their effect size. PRS are gaining popularity due to their potential to enable many applications such as testing genetic overlap between traits, enabling risk stratification, and serving as an aid in diagnosis prediction and personalised treatment (29). We use PRS for MDD, BMI, insomnia and headaches to test for evidence of non-specific or shared genetic factors underpinning antidepressant-specific side effects. Overall, our results suggest drug exposure alone does not explain the occurrence of side effects, and a combination of specific and non-specific factors underlie their aetiology.

## Methods

### Sample recruitment and genotyping

We use the Australian Genetics of Depression Study (AGDS), in which participants provide self-report responses on psychosocial factors of depression heterogeneity and antidepressant treatment outcomes (N= 20,941 with reported depression diagnosis) as well as DNA samples for genetic analysis. Sample recruitment has been described in detail elsewhere (30). Briefly, 14.3% of volunteers were recruited by mail invitations distributed by the Australian Department of Human Services (DHS) encouraging individuals who had previously used prescription antidepressants in the last 4.5 years to participate. Remaining participants were recruited through a nationwide media publicity campaign. This campaign targeted individuals who have sought medical attention by a psychiatrist or a psychologist for clinical depression. Recruited participants were directed to the study website to complete consent forms before answering the online surveys. Once the surveys were completed, and informed consent for donation of a DNA sample was given, a GeneFix GFX-02 DNA extraction kit (Isohelix plc) was sent to participants to collect 2mL of saliva for DNA extraction. Genotyping was performed using the Illumina Global Screening Array (GSA V.2.0.). Genotype data were cleaned by removing unknown or ambiguous map position, strand alignment, high missingness (>5%), deviation from Hardy-Weinberg equilibrium, low minor allele frequency (<1%) and GenTrain score <0.6 variants. Imputation was performed through the Michigan imputation server web service using the HRCr1.1 reference panel. Genotyped individuals were excluded from PRS analyses based on high genotype missingness, inconsistent and unresolvable sex or if deemed ancestry outliers from the European population, based on principal components derived from the 1000Genomes reference panel. The QIMR Berghofer Medical Research Institute Human Research Ethics Committee approved all questionnaires and research procedures.

### Phenotype ascertainment

This study focuses on participant-reported antidepressant adverse side effects. Participants first confirmed they had taken any of the ten most commonly prescribed antidepressants in Australia (sertraline, escitalopram, venlafaxine, fluoxetine, citalopram, desvenlafaxine, duloxetine, mirtazapine, amitriptyline and paroxetine). For each antidepressant taken, participants were asked whether they had experienced side effects and, if they did, to select which from a checklist with the twenty-three most commonly reported antidepressant side effects including *reduced sexual drive or desire, weight gain, dry mouth, nausea, drowsiness, insomnia, dizziness, fatigue, sweating, headache, suicidal thoughts, anxiety, agitation, shaking, constipation, diarrhoea, suicide attempt, blurred vision, muscle pain, vomiting, weight loss, runny nose* and *rash*.

### Side effect correlations and structural equation modelling

We used tetrachoric correlations as implemented in the psych library in R v3.6.1 to estimate the correlation (i.e. co-occurrence within the same set of people) of side effects across medications. Pairwise complete observations were used for these analyses. The correlation matrix was transformed into a distance matrix which was subjected to a minimum variance hierarchical clustering analysis using the *scipy* library in Python 3. The results are visualised with a clustergram generated using the *seaborn* and matplotlib libraries in Python 3.6. We further used structural equation modelling (OpenMx Rv3.6.2) to assess whether, for each side effect, there was evidence for drug class-specific factors over and above a common factor. For each side effect, we fit a bi-factor model consisting of a general factor loading onto the 10 binary side effects, and two drug-class factors “*SSRI*” and “*SNRI*” loading onto side effects from their respective drug class. All of the latent factors are orthogonal to each other. We refer to this model as the full model. Reduced models are also fit by removing the drug class factors one at a time; these are the SSRI and SNRI models (**Supplementary Figure 2**). Finally, a model consisting of a single general factor is also used for completeness. The four models are fit to the data using a full information maximum likelihood estimation assuming a liability threshold model for the binary manifest variables. After fitting, the simpler models are compared to the full model by the Akaike information criterion (AIC) and likelihood ratio test (LRT) with the mxCompare function implemented in OpenMx. Under this approach, the p-value represents whether a nested reduced model is losing a significant amount of information compared to its full counterpart. Thus, a statistically significant p-value indicates that removal of that drug class factor results in a poorer fit.

### Genetic instruments and polygenic risk scoring

To avoid biases due to population stratification and cryptic relatedness, only unrelated individuals of European ancestry were included in the genetic part of this study. Polygenic risk scores were calculated as a proxy for an individual’s genetic liability to a trait. In this study, we used publicly available GWAS results for depression(31), insomnia(32), chronic headaches, and BMI (from Benjamin Neale’s UK Biobank GWAS public repository). Genetic variant effect sizes were acquired from the GWAS data and used to calculate the predictive genetic risk for the traits investigated. Prior to estimating polygenic risk scores, we excluded low (r^2^ < 0.6) imputation quality and strand-ambiguous variants. We used two approaches to deal with correlation among genetic variants emerging through linkage disequilibrium (LD). First, we employed a recently developed powerful method named SBayesR (33). SBayesR estimates a conditional GWAS (i.e., one including all of the genetic variants as predictors at the same time) using marginal GWAS summary statistics and LD measures between genetic variants (LD matrix) under a Bayesian multiple regression framework. This method has been shown to improve polygenic prediction of complex traits. We also employed a more traditional clumping and thresholding procedure as sensitivity analyses. Briefly, PLINK (1.9) (34) was used to detect independent SNPs through a conservative clumping (p1 = 1, p2 = 1, r^2^ = 0.1, kb=10000) adjustment of linkage disequilibrium. Various p-value thresholds (p < 5 × 10-8, p < 1 × 10-5, p < 0.001, p < 0.01, p < 0.05, p < 0.1, p < 0.5, p < 1) were used to determine which variants to include for PRS calculation. Imputed genotype dosage data were used to calculate PRS by multiplying the variant effect size times the dosage of the effect allele. Finally, the total sum was calculated across all variants.

### PRS side effect association

Logistic regressions were used to examine the association between participant-reported side effects and PRS. The regressions were adjusted for sex, age at study enrollment and the first 20 genetic principal components to further adjust for potential population stratification. Variance explained was calculated as Nagelkerke’s pseudo R^2^:

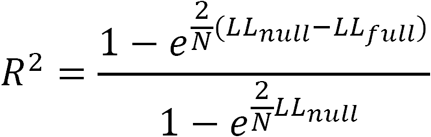

Where LL_full_ and LL_null_ are the log-likelihoods for the model with and without the PRS, respectively. Nominally significant results are defined as those with p<0.05, and statistical significance was defined after Bonferroni correction for multiple testing. For the MDD analysis, we adjusted for the association of MDD PRS with the 25 side effects across medications (p<0.002). For the other PRS, we adjusted for the testing of ten drugs across (p<0.005), and for the sensitivity analyses (using clumping and thresholding), we adjusted for eight thresholds times ten medications (p<0.000625). This method is rather conservative, as it does not account for the moderate to high co-linearity within the eight PRS and within the side effects across medications. Results are visualised as heatmaps of variance explained using *seaborn* and *matplotlib* in Python 3.6. These analyses were performed using complete case data.

## Results

### Demographics and side effect prevalence

As previously reported (30), the majority of AGDS participants were female (75%). The average age was 43 (s.d.=15.3) years old. Most people (60-75%) reported at least one side effect regardless of antidepressant taken (**Supplementary Figure 1**). **Figure 1** shows the prevalence of reported side effects for males and females across the ten studied antidepressants. *Reduced sex drive* and *weight gain* had the highest prevalence. The least prevalent side effects were *suicide attempt, blurred vision, rash, weight loss*, and *muscular pain*. Overall, there were significant differences between the prevalence of side effects for males and females across all medications (**Table 1** and **Supplementary Table S1**). Males reported experiencing *reduced sex drive or function* more often than females. Conversely, women were more likely to report *weight gain* compared to males (**Supplementary Table S1**). Other side effects showing a robust difference (i.e., a difference observed for five or more medications) in prevalence between males and females included nausea, *headaches, dizziness, shakes* and *vomits*.

**Table 1.** AGDS demographics and side effect prevalence across medications

|  |  |  |  |  |  |  |
| --- | --- | --- | --- | --- | --- | --- |
|  | Males N | Females N | Sex p-value<sup>¥</sup> | Endorsed side effect age (s.d.) | Not endorsed side effect age (s.d.) | Age p-value<sup>§</sup> |
| **N** | 5111 | 15830 | - | - | - | - |
| **Age mean (s.d.)** | 47.99 (15) | 41.41 (14) | 6.6e-160 | - | - | - |
| **Reduced sexual desire** | 2251 (44%) | 6264 (39%) | 1.5e-08 | 41.0 (13.98) | 44.4 (16.01) | 1.1e-58 |
| **Weight gain** | 1402 (27%) | 5695 (35%) | 3.2e-29 | 41.9 (13.89) | 43.6 (15.96) | 1.1e-13 |
| **Dry mouth** | 1236 (24%) | 4544 (28%) | 3.2e-10 | 42.7 (14.20) | 43.1 (15.71) | 0.071 |
| **Nausea** | 867 (16%) | 4352 (27%) | 1e-51 | 37.7 (13.34) | 44.8 (15.52) | 1.6e-187 |
| **Headaches** | 704 (13%) | 3282 (20%) | 3.1e-28 | 38.3 (13.74) | 44.1 (15.45) | 2e-106 |
| **Dizziness** | 959 (18%) | 3930 (24%) | 5.2e-19 | 38.4 (13.43) | 44.4 (15.57) | 5.3e-130 |
| **Shakes** | 571 (11%) | 2466 (15%) | 7.4e-15 | 38.8 (14.22) | 43.7 (15.37) | 3.8e-62 |
| **Muscle pain** | 234 (4%) | 837 (5%) | 0.045 | 42.0 (14.98) | 43.1 (15.33) | 0.029 |
| **Sweating** | 997 (19%) | 3291 (20%) | 0.048 | 40.3 (13.75) | 43.7 (15.61) | 6.8e-38 |
| **Vomit** | 147 (2%) | 826 (5%) | 4.7e-12 | 35.7 (12.65) | 43.4 (15.34) | 6e-53 |
| **Constipation** | 395 (7%) | 1489 (9%) | 2.70E-04 | 43.3 (15.01) | 43.0 (15.34) | 0.440 |
| **Diarrhoea** | 368 (7%) | 1176 (7%) | 0.590 | 39.9 (13.87) | 43.3 (15.39) | 8.8e-17 |
| **Drowsiness** | 1173 (22%) | 3709 (23%) | 0.480 | 39.7 (14.30) | 44.0 (15.46) | 6.8e-67 |
| **Trouble sleeping** | 1052 (20%) | 3672 (23%) | 1.00E-04 | 39.6 (14.35) | 44.0 (15.43) | 2.3e-70 |
| **Anxiety** | 794 (15%) | 2973 (18%) | 1.5e-07 | 39.1 (14.24) | 43.9 (15.40) | 1.6e-68 |
| **Agitation** | 786 (15%) | 2816 (17%) | 7.2e-05 | 39.6 (14.12) | 43.7 (15.45) | 1.3e-50 |
| **Fatigue** | 912 (17%) | 3181 (20%) | 4.20E-04 | 39.4 (14.49) | 43.9 (15.38) | 8.4e-63 |
| **Weight loss** | 157 (3%) | 795 (5%) | 5.9e-09 | 36.1 (13.42) | 43.3 (15.32) | 5.1e-47 |
| **Rashes** | 97 (1%) | 257 (1%) | 0.190 | 45.5 (14.96) | 43.0 (15.31) | 0.002 |
| **Runny nose** | 82 (1%) | 344 (2%) | 0.012 | 44.9 (15.64) | 43.0 (15.30) | 0.010 |
| **Blurry vision** | 266 (5%) | 1012 (6%) | 0.002 | 42.6 (14.60) | 43.0 (15.35) | 0.280 |
| **Suicide thoughts** | 699 (13%) | 2560 (16%) | 1.9e-05 | 38.2 (14.24) | 43.9 (15.33) | 4.7e-87 |
| **Suicide attempt** | 248 (4%) | 1090 (6%) | 2.4e-07 | 35.9 (13.41) | 43.5 (15.31) | 4.5e-69 |
| **Other side effects** | 632 (12%) | 1968 (12%) | 0.900 | 41.3 (14.05) | 43.3 (15.47) | 1.3e-09 |
| **No side effects** | 334 (6%) | 1140 (7%) | 0.110 | 43.8 (15.04) | 43.0 (15.33) | 0.044 |
| <sup>¥</sup>Two sample Z proportion test. <sup>§</sup>Two sample t-test. Data of each side effect per medication studied is available in Supplementary Table 1 |  |  |  |  |  |  |

**Figure 1.**
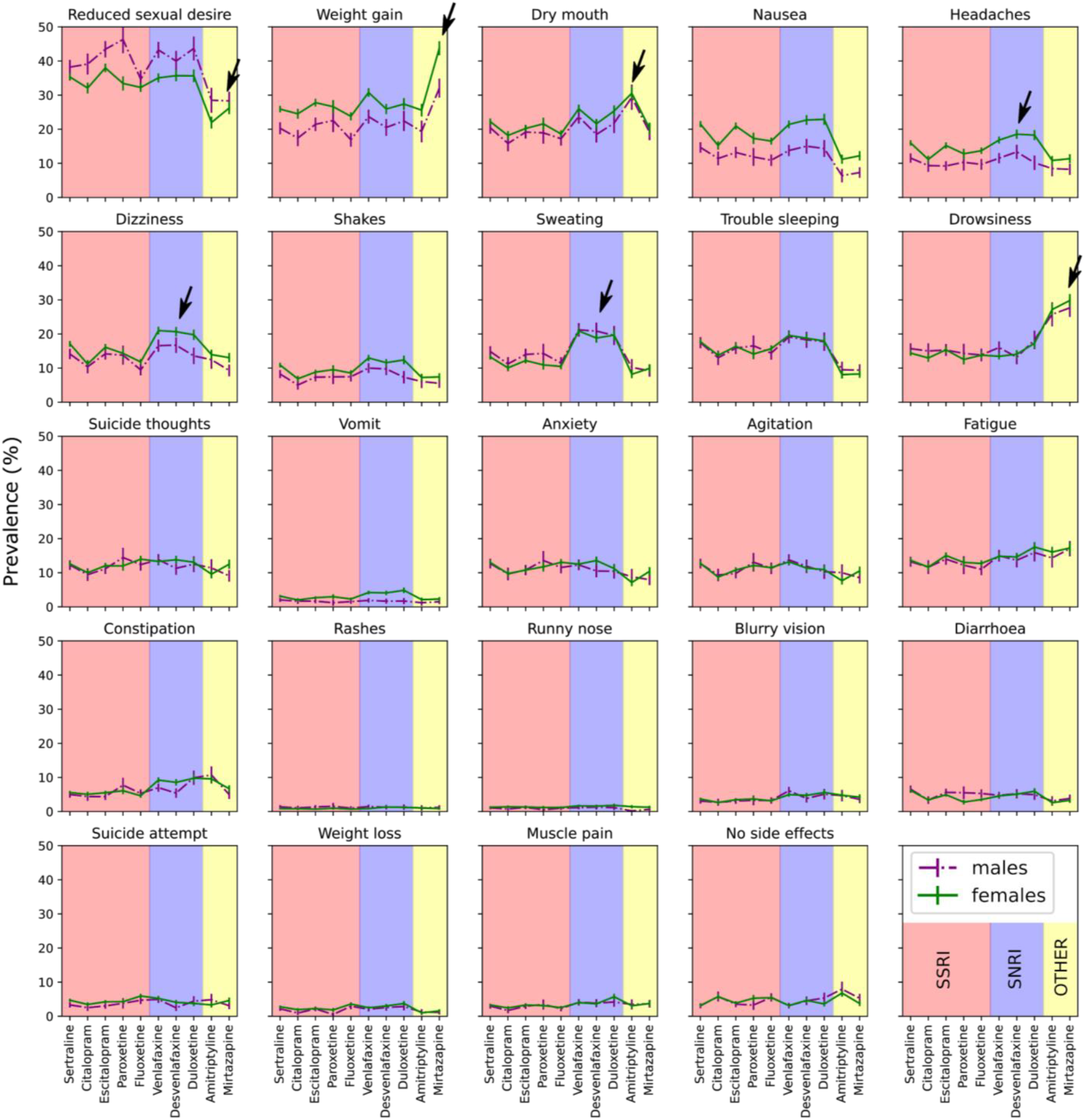
Side effect prevalence across medications. Plots depicting the prevalence and 95% confidence interval for each of the 25 studied side effects across ten medications. Results are split by males (dotted lines) and females (solid lines). The plots are shaded according to antidepressant drug class. Some of the findings discussed in the results are highlighted by black arrows. P values are available in **Supplementary Table S1**.

Overall, participants reporting specific side effects were younger than those not doing so. Conversely, two side effects, *rashes* and *runny nose* were reported by older participants (**Table 1**). Some side effects such as *dizziness, headaches* and *sweating* showed a higher prevalence across SNRIs compared to SSRIs. These patterns were sometimes sex-specific (**Figure 1** and **Supplementary Table S1**). We observed a higher prevalence of *weight gain* from mirtazapine, *dry mouth* from amitriptyline and *drowsiness* from both amitriptyline and mirtazapine than SNRIs and SSRIs. Conversely, *reduced sexual desire or function* was more commonly reported for SSRIs or SNRIs compared to amitriptyline and mirtazapine. Finally, side effects such as *sweating, headaches* and *dizziness* were reported more often for SNRIs (**Figure 1**).

### Side effects co-occur across medications

For each antidepressant taken, participants reported whether or not they had experienced specific side effects. Some individuals had taken more than one antidepressant. Thus, several subsets of participants with overlapping data for different antidepressants were available. This overlap enabled us to assess whether the same side effects occur independently across different antidepressants or whether they co-occur (covary) across medications. We identified high correlations for side effects across medications. A clustering based on these correlations grouped these variables by side effect rather than by antidepressant or medication class (**Figure 2**). Overall, these results would suggest that antidepressant side effects co-occur in the same people regardless of the medication of choice or their class, which would be consistent with the existence of a set of general non-specific (i.e. independent of drug mode of action) factors partly underlying their aetiology. Structural equation models were used to compare a bifactor model with both common and drug-class factors against reduced models, including only specific drug classes or no drug class (**Supplementary Figure 2a**). Our results suggested that the common factor captured a high proportion of the covariance across medications. A single common factor model was preferred over the SSRI, SNRI, or the full model for some side effects. However, we identified evidence for medication-class specific effects for some side effects. For example, *sweating* and *suicide thoughts* showed evidence of an SNRI-specific factor, and *vomits* and *shakes* showed evidence of an SSRI-specific factor. In contrast, dizziness and *constipation* showed evidence for both factors over and above a general factor (**Supplementary Figure 2b** and **Supplementary Table S2**).

**Figure 2.**
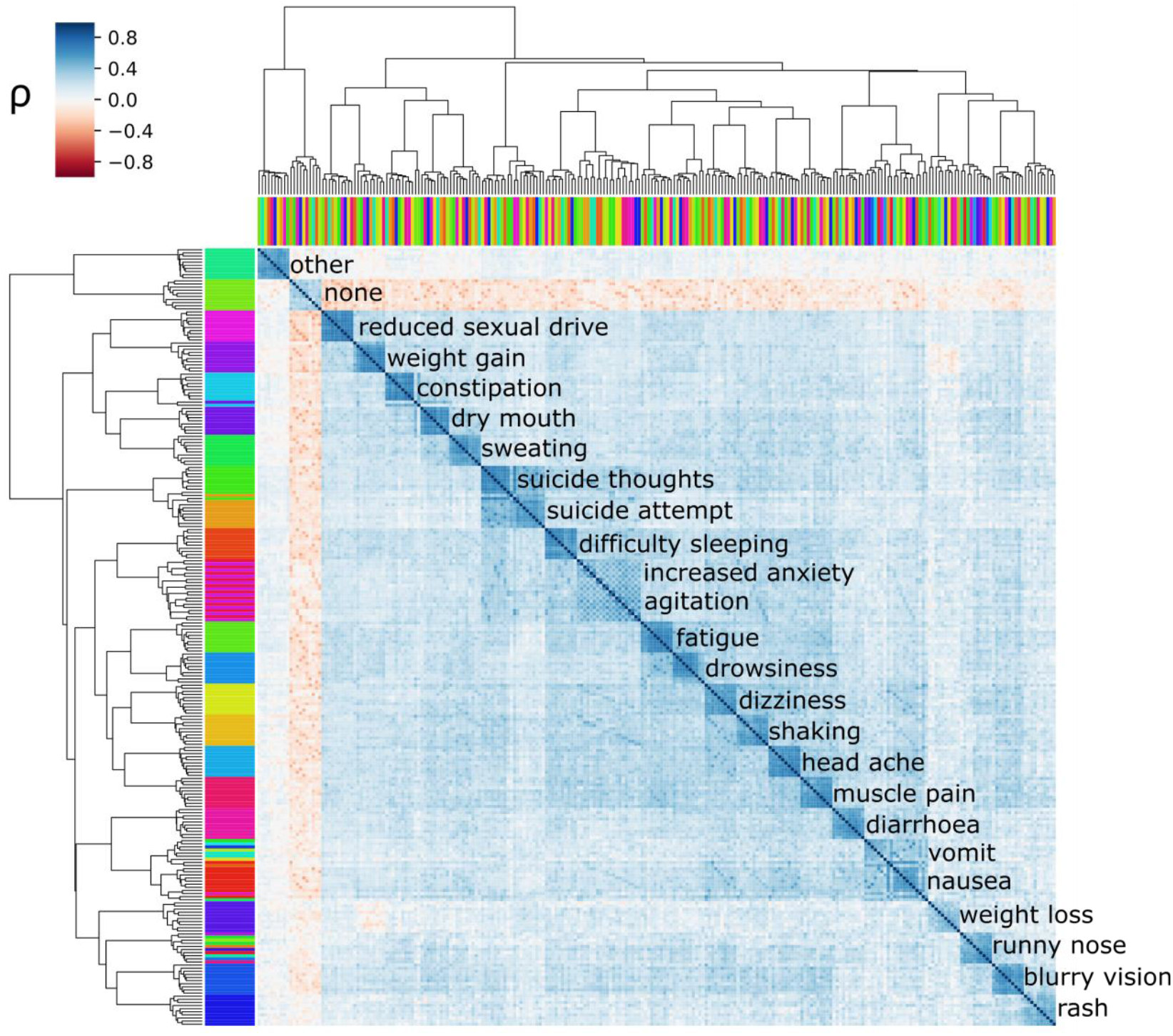
side effects co-occur across medications. Results of hierarchical clustering based on tetrachoric correlations of the side effects across drugs. The colour shows the amount of overlap (co-occurrence) of side effects of patient-reported side effects. The top and side colour bars represent the ten medications and the 25 studied side effects, respectively. Labels show the most abundant side effect in each cluster. ρ-tetrachoric correlation coefficient.

### Genetic factors underlie side effects

We employed a polygenic risk scoring strategy to assess whether the general factors identified have a genetic basis (**Figure 3a**; see methods). Depression PRS were positively associated with most side effects when pooling data across medications. The side effects that showed the strongest association with depression PRS were suicide thoughts and suicide attempt, which is consistent with their intricate relationship with depression (**Figure 3b**; **Table 2**). The strength of these associations was reduced, and increased heterogeneity was observed when splitting per antidepressant regardless of what PRS method was used (**Supplementary Figure 3**). Only dizziness, constipation, agitation, suicide thoughts, suicide attempt and reduced sexual desire showed significant association with depression PRS when splitting per antidepressant (**Supplementary Figure 3; Supplementary Table S3**).

**Table 2.** Polygenic prediction of specific side effects across medications

| Antidepressant | BMI PRS predicting weight gain |  |  | Insomnia PRS predicting trouble sleeping |  |  | Headaches PRS predicting headaches |  |  |
| --- | --- | --- | --- | --- | --- | --- | --- | --- | --- |
|  | OR (95%C.I.) | P-value | Variance explained (%) | OR (95%C.I.) | P-value | Variance explained (%) | OR (95%C.I.) | P-value | Variance explained (%) |
| Sertraline | 1.24 (1.17—1.32) | **4.66E-12** | 1.25 | 1.06 (0.99—1.14) | 0.077 | 0.09 | 1.16 (1.08—1.25) | **8.17E-05** | 0.49 |
| Escitalopram | 1.21 (1.13—1.29) | **3.58E-08** | 1.01 | 1.12 (1.03—1.21) | 0.008 | 0.27 | 1.06 (0.97—1.16) | 0.178 | 0.08 |
| Venlafaxine | 1.20 (1.12—1.28) | **3.72E-07** | 0.93 | 1.03 (0.95—1.11) | 0.463 | 0.02 | 1.11 (1.01—1.21) | 0.025 | 0.22 |
| Amitriptyline | 1.25 (1.12—1.40) | **9.84E-05** | 1.38 | 1.28 (1.07—1.52) | 0.007 | 1.03 | 1.09 (0.93—1.28) | 0.268 | 0.15 |
| Mirtazapine | 1.20 (1.09—1.32) | **2.03E-04** | 0.95 | 1.03 (0.88—1.20) | 0.745 | 0.01 | 1.04 (0.90—1.20) | 0.560 | 0.04 |
| Desvenlafaxine | 1.22 (1.11—1.33) | **3.01E-05** | 1.03 | 1.13 (1.03—1.25) | 0.015 | 0.38 | 1.08 (0.98—1.20) | 0.126 | 0.16 |
| Citalopram | 1.37 (1.24—1.50) | **9.06E-11** | 2.5 | 1.08 (0.96—1.21) | 0.194 | 0.12 | 1.11 (0.98—1.26) | 0.088 | 0.23 |
| Fluoxetine | 1.26 (1.17—1.36) | **5.75E-09** | 1.42 | 1.03 (0.94—1.13) | 0.523 | 0.02 | 1.02 (0.93—1.12) | 0.689 | 0.01 |
| Duloxetine | 1.20 (1.08—1.32) | **3.60E-04** | 0.94 | 1.14 (1.02—1.28) | 0.026 | 0.4 | 1.03 (0.92—1.15) | 0.626 | 0.02 |
| Paroxetine | 1.22 (1.09—1.36) | **6.28E-04** | 1.1 | 1.16 (1.01—1.33) | 0.042 | 0.47 | 1.16 (1.00—1.35) | 0.053 | 0.46 |
| Results from logistic regressions predicting weight gain, trouble sleeping and headaches using BMI, Insomnia and headaches PRS respectively. Bolded p-values represent values significant after multiple testing correction (p<0.005). Results shown for SBayesR, for clumping and thresholding sensitivity results see Supplementary Tables. |  |  |  |  |  |  |  |  |  |

**Figure 3.**
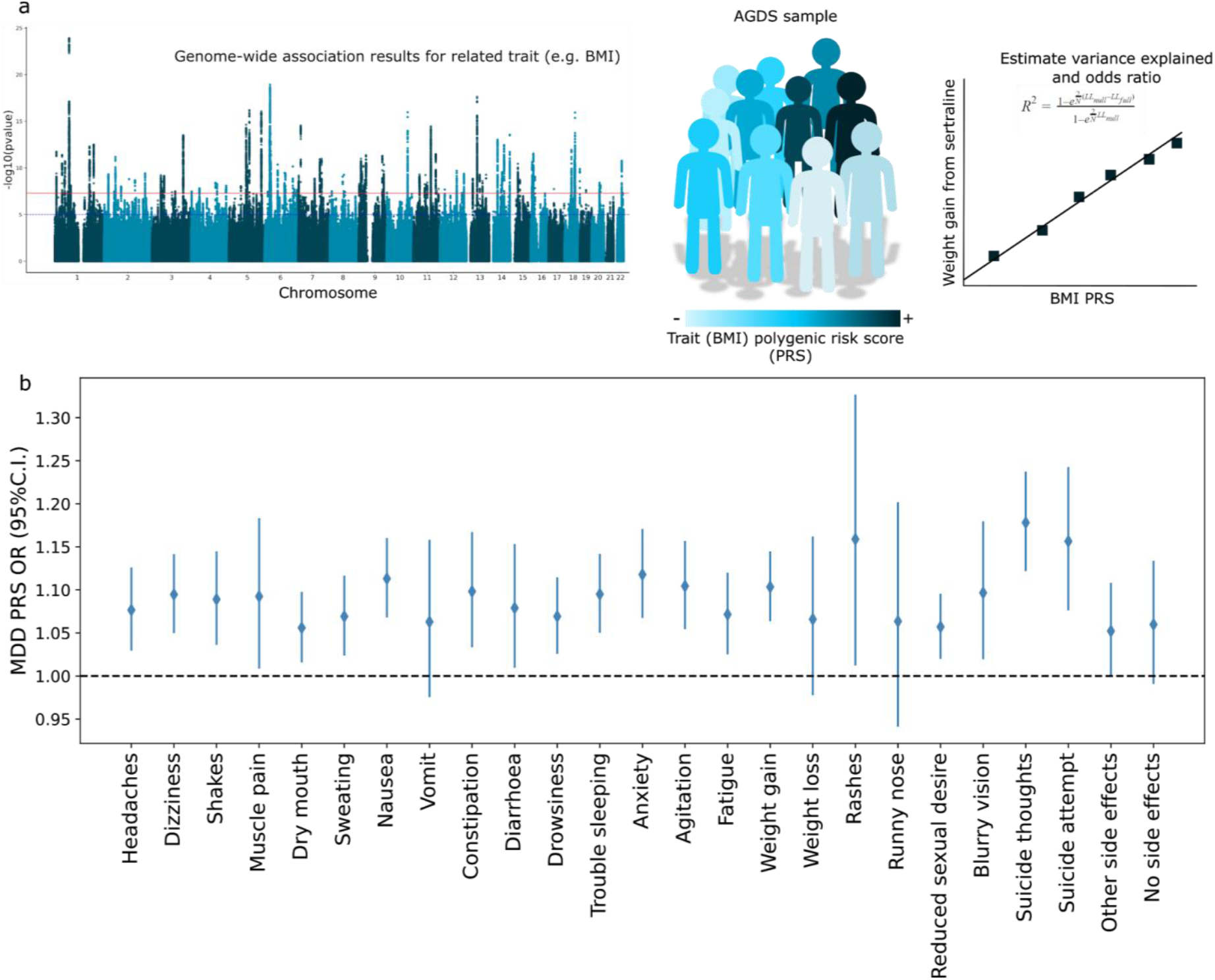
Genetic factors underlying side effects. a) Overview of the polygenic risk scoring approach employed. Results from a published GWAS that do not include the AGDS sample are used to estimate the polygenic risk for a given trait in the AGDS. These scores can be used to predict the side effect of interest.b) Forest plot showing the results (odds ratio and 95% confidence intervals) for the association between MDD PRS and side effects across antidepressants.

We then tested whether polygenic risk for related common traits underlies the risk for specific side effects. We chose to study weight gain, insomnia and headaches as we could identify related complex traits (BMI, insomnia and headaches, respectively) for which GWAS data is available. BMI PRS were strongly and robustly associated with weight gain across all medications (**Figure 4; Supplementary Table S4**). PRS for headaches and insomnia showed evidence of association with headaches and trouble sleeping as side effects. PRS for headaches were associated with headaches from sertraline and, to a lesser extent, with headaches from venlafaxine. Insomnia PRS showed suggestive evidence of predicting insomnia from escitalopram and amitriptyline (**Supplementary Figure 4; Supplementary Tables S5-6**). Amitriptyline is sometimes used to treat insomnia. In fact, we identified a higher insomnia PRS on average for participants that reported taking amitriptyline (**Supplementary Figure 5**). To assess whether this could be confounding the association, we repeated the analysis adjusting for whether participants reported taking amitriptyline for insomnia. Insomnia PRS was associated with participants taking amitriptyline for insomnia (OR=1.2 95%C.I.=[1.1-1.3]). Nonetheless, whether participants took amitriptyline for insomnia was not associated with insomnia as a side effect from this medication (OR=1.2 95%C.I.=[0.8-1.8]). As expected, insomnia PRS was still predictive of insomnia as a side effect from amitriptyline after adjusting for whether it was taken for insomnia or not (OR=1.3 95%C.I.=[1.1-1.5]). Overall, the direction of effects between PRS and side effects were positive for all medications (e.g., higher BMI PRS higher risk of weight gain as a side effect). Still, the errors varied potentially due to sample size differences.

**Figure 4.**
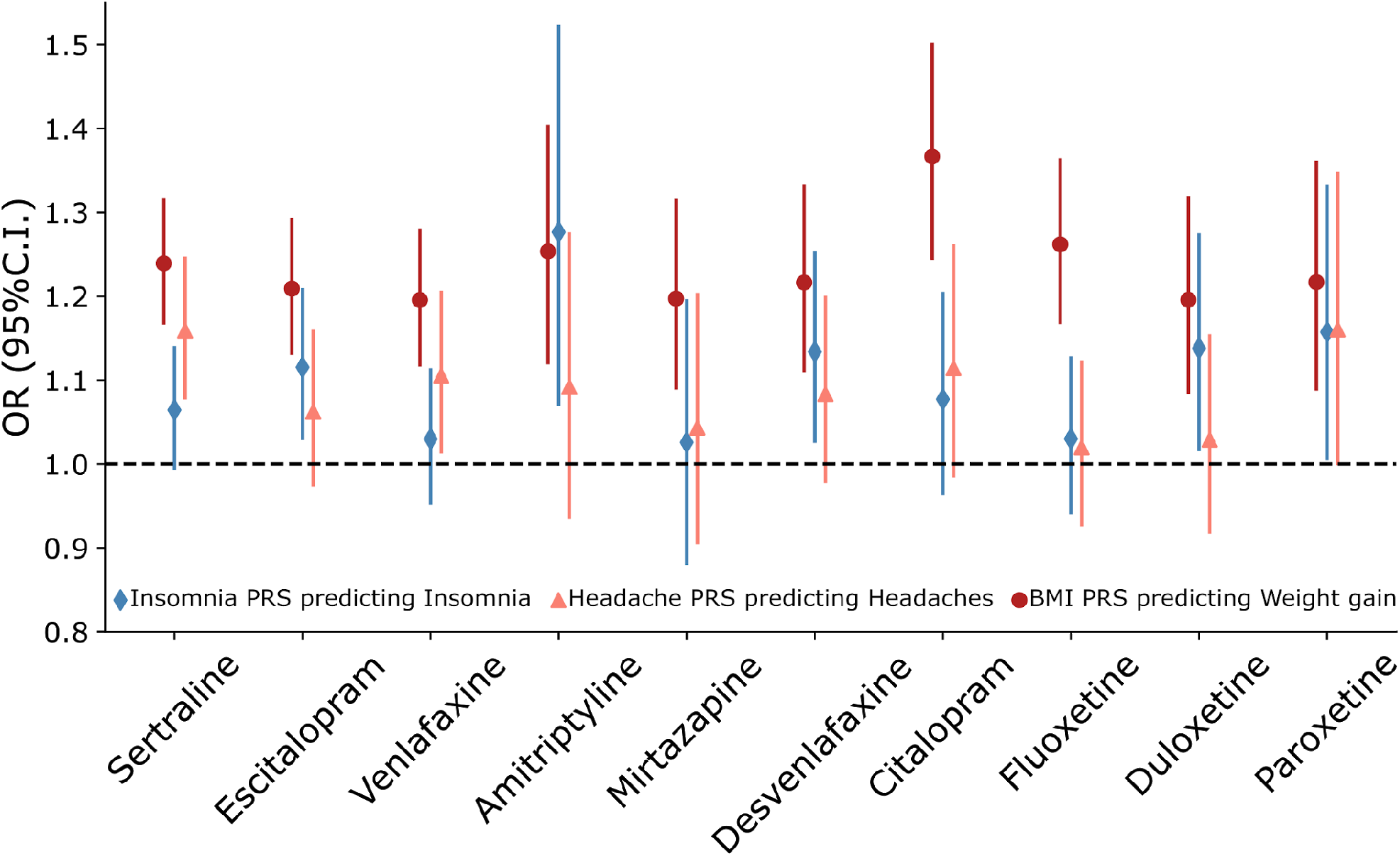
Genetic prediction of antidepressant side effects. Forest plots show the results of associations between polygenic risk scores for BMI, insomnia and chronic headaches predicting weight gain, insomnia and headaches reported as side effects for antidepressant medications. Markers represent odds ratios, and error bars represent 95% confidence intervals.

## Discussion

In this study, we aimed at gaining insights into the genetic aetiology of self-reported antidepressant adverse side effects. We identified the most common side effect to be *reduced sexual drive or function*, followed by *weight gain. Reduced sexual function* was most prevalent among males taking paroxetine, an SSRI, whereas *weight gain* was the most prevalent among females taking amitriptyline, a tricyclic antidepressant (TCA). These findings are consistent with research findings showing SSRIs exhibit the most adverse sexual effects (35), whereas TCAs have been established to cause weight increases (36).

Studies have suggested that SSRI-based sexual dysfunction may result in 40% to 65% of individuals ceasing treatment (37). It is hypothesised that testosterone and dopamine neurotransmitters are dysregulated by SSRIs, a plausible hypothesis considering the role that testosterone plays in sexual function and the high amounts that circulate in males compared to females (38,39). Moreover, serotonin plays a crucial role in initiating smooth muscle contraction of the genito-urinary system and regulating the response of the sexual cycle; thus, exogenous substances such as SSRIs that alter these mechanisms may cause sexual dysfunction in both males and females (40). SNRIs have been reported to lead to more *dizziness, trouble sleeping* and *dry mouth* than SSRIs (36,41). We found a higher prevalence for dizziness and sweating from venlafaxine, desvenlafaxine and duloxetine (SNRIs) compared to other antidepressants.

SSRIs are generally prescribed as first-line agents due to their safety profile, evident since clinical studies from the 1980s onwards, for individuals with multiple comorbidities (42–44). Moreover, as they are better tolerated than older agents, this results in more effective long-term management (45). While we observed specific instances where side effects were more prevalent across other antidepressant types, we did not find a lower prevalence for *any side effect* nor a higher prevalence of *no side effects* for SSRIs. This might be explained by the fact that nonspecific factors (i.e., regardless of medication class) seemed to underlie the self-reporting of side effects.

Side effects co-occurred across all antidepressants assessed despite their variable modes of action and metabolism (46). Participants who reported a side effect for one medication were more likely to report that same side effect for other medications. This strongly suggests the existence of common non-specific factors, which are potentially a mixture of shared pharmacological and genetic factors, underlying their aetiology. Our structural equation modelling analyses suggested that for some side effects, a common factor model would be preferred over models considering the drug classes.

The evidence for a lack of drug class factors was not equally strong for all side effects. For example, *sweating* showed the best fit to a model containing an SNRI factor along with the common factor. This is consistent with the higher prevalence of sweating reported for SNRIs compared to other drugs, which would already imply an SNRI-specific factor increasing its prevalence. For some side effects, the existence of common factors is further supported by the fact that they are widely reported from other types of medications; for example, *weight gain* is a common side effect not only for antidepressants but also for antipsychotics, antihyperglycemics, antihypertensives and corticosteroids (47). The nature of these common factors is complex and might include a mixture of a *nocebo* effect (48), shared metabolism, common environmental and genetic factors.

This study uniquely addressed the role of genetic factors on antidepressant adverse side effects. First, we evaluated whether the genetic liability to depression, which has been linked to increased depression severity, recurrence and persistence (49), is associated with antidepressant side effects. Depression PRS were associated with many side effects, particularly those that could be considered depression symptoms or common comorbidities such as anxiety, trouble sleeping and suicidal behaviours. When testing across individual antidepressants, a more heterogeneous pattern was observed. Although we cannot rule out reduced power due to subsampling when performing these analyses, the heterogeneity could imply that interactions between specific drugs and depression PRS underlie the studied adverse side effects. For example, MDD PRS was robustly associated with increased *suicidality* for venlafaxine, but evidence for association with other drugs did not reach statistical significance. This result is not easily attributed to power given that sertraline, and not venlafaxine, is the drug for which we have the largest sample size. Furthermore, venlafaxine has been associated with increased suicidality compared to placebo in modern meta-analyses (50,51). This is also consistent with our findings of an SNRI factor underlying suicide thoughts over and above a general factor. Our results suggest that this increased risk might be mediated by genetic factors, which opens up the opportunity for genetic risk stratification, management and genetically informed therapies.

The second type of factor consisted of liability to traits related to the nature of specific side effects. We tested for the association between PRS for BMI, chronic headaches and insomnia with *weight gain, headaches* and *trouble sleeping* as side effects from antidepressants. We found strong evidence for BMI PRS predicting *weight gain* for all medications. Evidence for *headaches* and *insomnia* was moderate, and associations remained heterogeneous across medications. These observations are consistent with *weight gain* being a potentially less specific side effect. As discussed above, weight gain is frequently reported for very distinct classes of drugs. On the other hand, it is unclear whether the heterogeneity observed for headaches and insomnia is due to power. The fact that the significant association for headaches was under sertraline, which is the most commonly reported drug in our sample, would be consistent with a lack of power underlying the lack of significance for other similar (SSRI and SNRI) pharmacological medications. Genetic risk for insomnia was associated with *insomnia* from amitriptyline. TCAs are generally associated with drowsiness and fatigue, but people with a higher genetic risk for insomnia were more likely to report insomnia as a side effect from amitriptyline. This may represent a prescribing bias caused by the fact that TCAs are often used (in lower doses) to treat insomnia. We performed a sensitivity analysis by adjusting for whether participants reported taking amitriptyline for insomnia and showed that this was not confounding the association we observed. Altogether, our results would suggest that in addition to genetic factors associated with depression, the genetic liability to side effect-related traits, such as BMI for weight gain, also underlie their aetiology. These results further prove the principle of using genetic data to study and predict antidepressant side effects. Genetics-driven prediction of treatment outcomes is one of the major challenges towards achieving precision medicine.

This study represents one of the most powered and comprehensive explorations of antidepressant side effects. The unprecedented detail and sample size of the AGDS enable us to gain valuable insights into the genetic and pharmacological underpinnings of antidepressant side effects. Nonetheless, certain limitations need to be acknowledged. First, the retrospective nature of this study and the reported phenotypes are prone to recall bias and subjective definition of a side effect. Second, side effect prevalence were estimated across the whole sample, including participants that have taken more than one medication. As such, the non-independence between these estimates should be taken into account when comparing side effect prevalence across medications. We did not collect information on the antidepressant dosages nor on more serious, albeit rarer, side effects such as the onset of mania, attempted suicide resulting in hospitalisation or myocardial infarction. Our genetic analyses were performed on a subset of individuals of European ancestry to prevent spurious associations arising from population stratification. Thus, caution must be taken when generalising our PRS findings to populations of a distinct genetic background.

In conclusion, we characterised the aetiology of side effects in a sample of Australian adults who reported depression over their lifetime. *Sexual dysfunction* and *weight gain* are the most commonly reported side effects. Some side effects, including weight gain and sexual dysfunction, showed clear differential sex-specific prevalences. We used clustering and structural equation modelling to test for co-occurrence and drug-class specificity of the side effects. We observed that side effects significantly co-occurred across medications, suggesting that shared pharmacological or genetic factors underlie their aetiology. As such, these reported side effects may be manifestations of depression severity, persistence, recurrence or non-response to treatment or its comorbidities. We employed PRS to test whether these shared factors had a genetic component. Depression PRS were associated with most of the side effects studied, strongly suggesting that a depression severity factor underpins reporting of some side effects. Furthermore, trait-specific PRS, such as BMI, were predictive of related side effects such as weight gain. Altogether our results suggest that drug exposure alone does not explain the occurrence of side effects, and a combination of specific and non-specific factors underlie their aetiology. Future studies on the genetic aetiology (e.g. performing well-powered GWAS) of adverse-side effects will enable further insights into their underpinnings as well as the possibility of genetically driven risk stratification and adverse side effect prophylaxis strategies.

## Data Availability

Summary data on prevalence and effects described in this manuscript is available in the supplementary data. GWAS summary statistics used in this study are publicly available and have been disclosed but are also available upon request. Code related to this study data analysis is available upon request from the authors. Access to the AGDS data is restricted due to the ethical guidelines governing the study.

## Acknowledgements

Data collection for AGDS was possible thanks to funding from the Australian National Health & Medical Research Council (NHMRC) to NGM, NRW, SEM, IHB, EMB, PAL (GNT1086683) and Medical Research Future Fund (APP1200644). We thank our colleagues Richard Parker, Simone Cross, Scott Gordon and Lenore Sullivan for their valuable work coordinating all the administrative and operational aspects of the AGDS project. AIC is supported by a UQ Research Training Scholarship from The University of Queensland (UQ). MER thanks the support of NHMRC and the Australian Research Council (ARC) through an NHMRC-ARC Dementia Research Development Fellowship (GNT1102821). SEM is supported in part by NHMRC investigator grant APP1172917. The views expressed are those of the authors and not necessarily those of the affiliated or funding institutions.

## Author contributions

AIC, NGM, SEM, NRW, IBH, EMB and MER designed this study. AIC performed the analyses. AIC and AM wrote the first version of the manuscript. AIC and JGT performed the SEM analyses. NGM, SEM, NRW, EMB, PAL and IBH designed and directed the AGDS data collection efforts. All authors contributed to the interpretation of the results and provided feedback on the preliminary versions of the manuscript.

## Competing interests

IBH has been: Commissioner of Australia’s National Mental Health Commission (2012–2018); Co-director of Health & Policy at the Brain & Mind Centre, University of Sydney; leading community-based and pharmaceutical industry-supported projects (Wyeth, Eli Lilly, Servier, Pfizer, AstraZeneca) focused on the identification and better management of anxiety and depression; a member of the Medical Advisory Panel for Medibank Private until October 2017; a board member of Psychosis Australia Trust; a member of the Veterans Mental Health Clinical Reference Group; and Chief Scientific Advisor to and an equity shareholder in Innowell. AIC, AM, NGM, JGT, PAL, SEM,PL, JGT, NRW, EMB and MER have nothing to disclose.

## Supplementary Materials

### Supplementary Table list (available as a worksheet)

S1 Prevalence of side effects across medications and demographic factors

S2 Fit statistics for structural equation models

S3 Association results between depression PRS and all side effects across medications

S4 Association results between BMI PRS and weight gain as a side effect across medications

S5 Association results between chronic headaches PRS and headaches as a side effect across medications

S6 Association results between Insomnia PRS and trouble sleeping as a side effect across medications

### Supplementary Figures

**Supplementary Figure 1.**
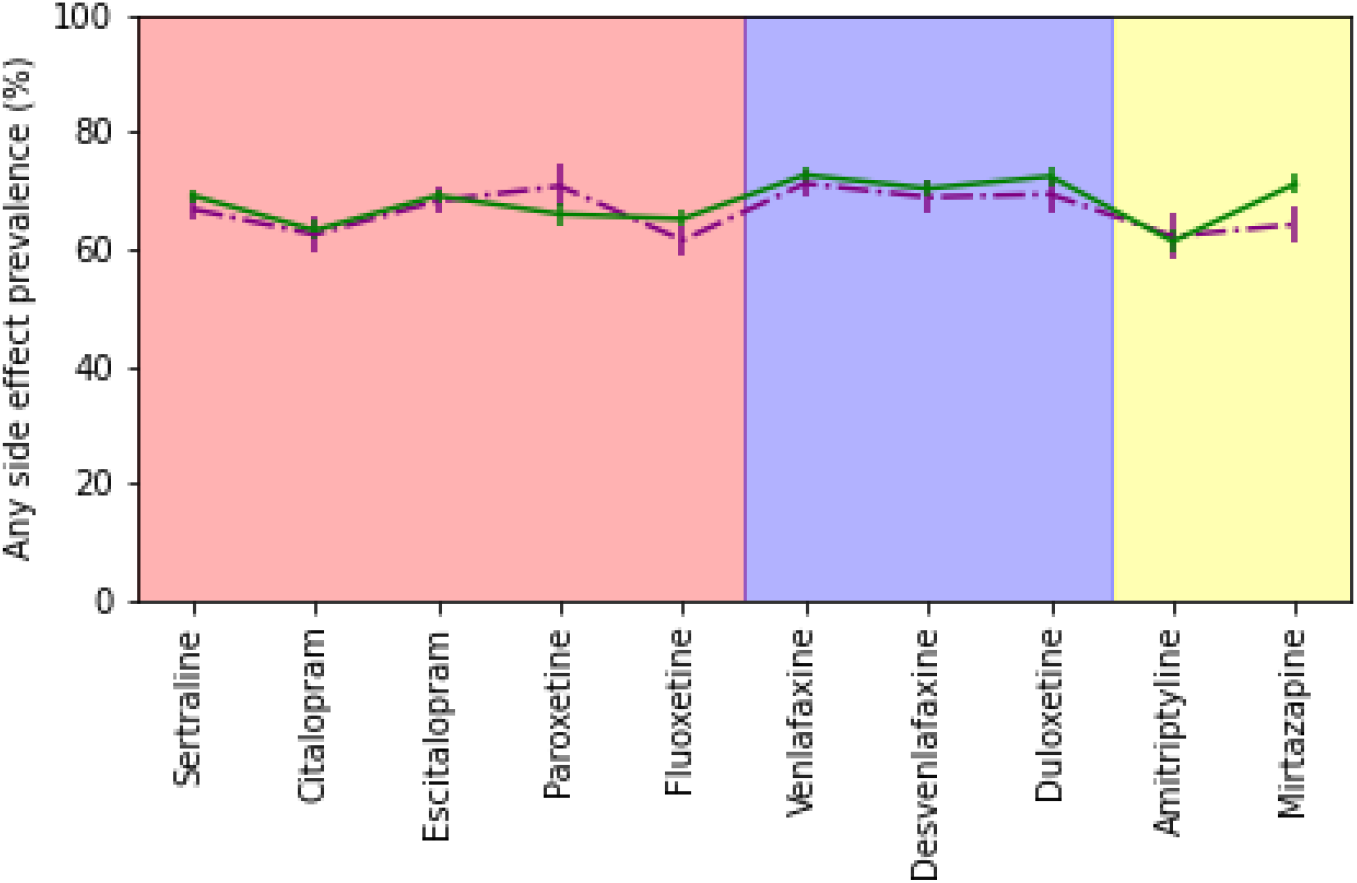
Prevalence of any side effect across medications. Plot depicting the prevalence and 95% confidence interval for any side effects across the ten medications under study.

**Supplementary Figure 2.**
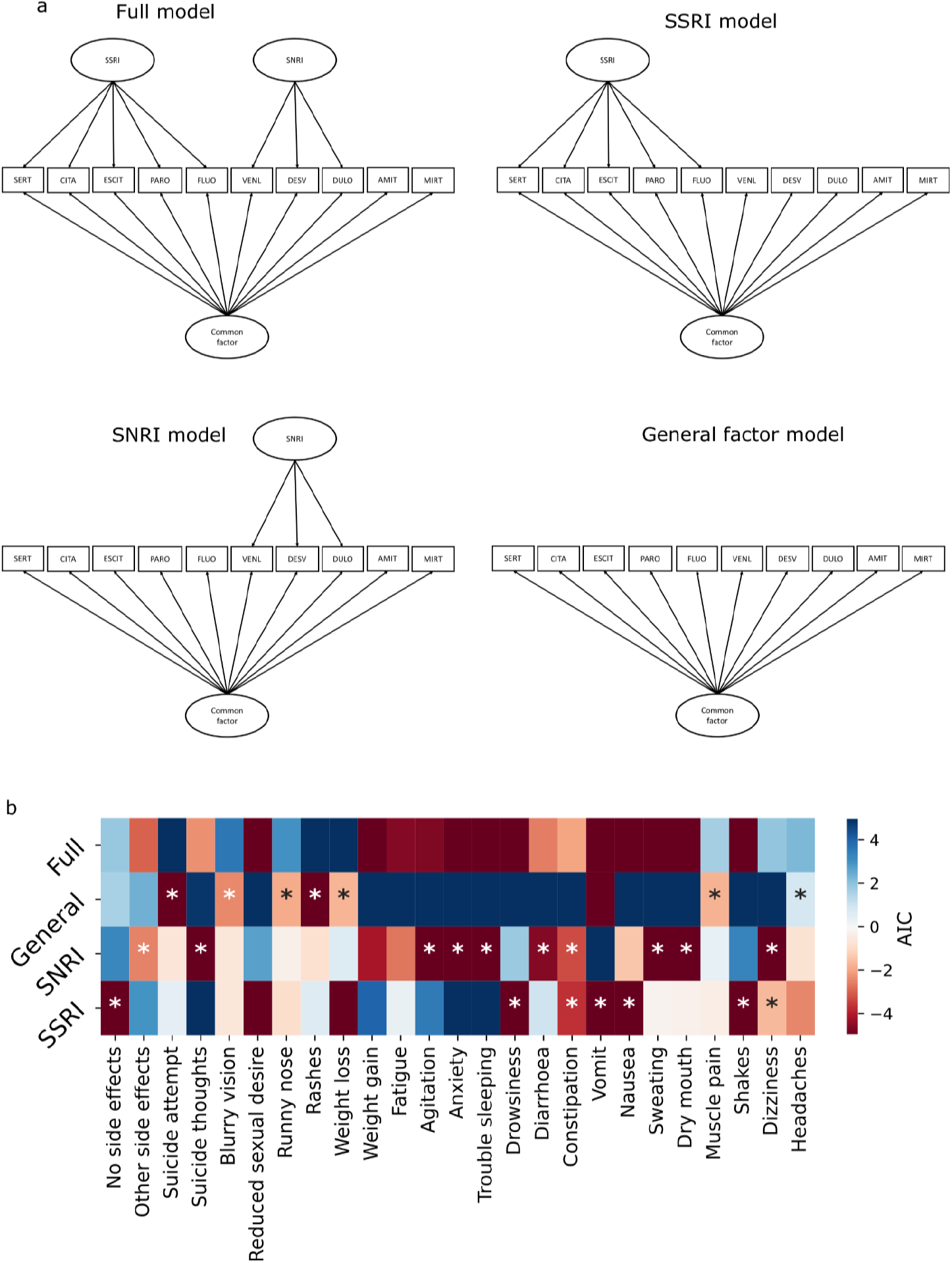
SEM analysis results. **a)** Path diagrams of the structural equation models fit via full information maximum likelihood using OpenMx. The manifest variables represent a reported side effect for each medication. b) Heatmap comparing the models Akaike information criterion (AIC). AIC values were mean-centred to keep them on the same scale; lower values represent better model fit. Stars mark the reduced model which would be chosen over the full model given their likelihood ratio test p-value. When two models are to be chosen, the most parsimonious one (if any) is preferred. **Supplementary Table 2** contains the relevant AIC values and likelihood ratio test p-values as well as the preferred model by each approach.

**Supplementary Figure 3.**
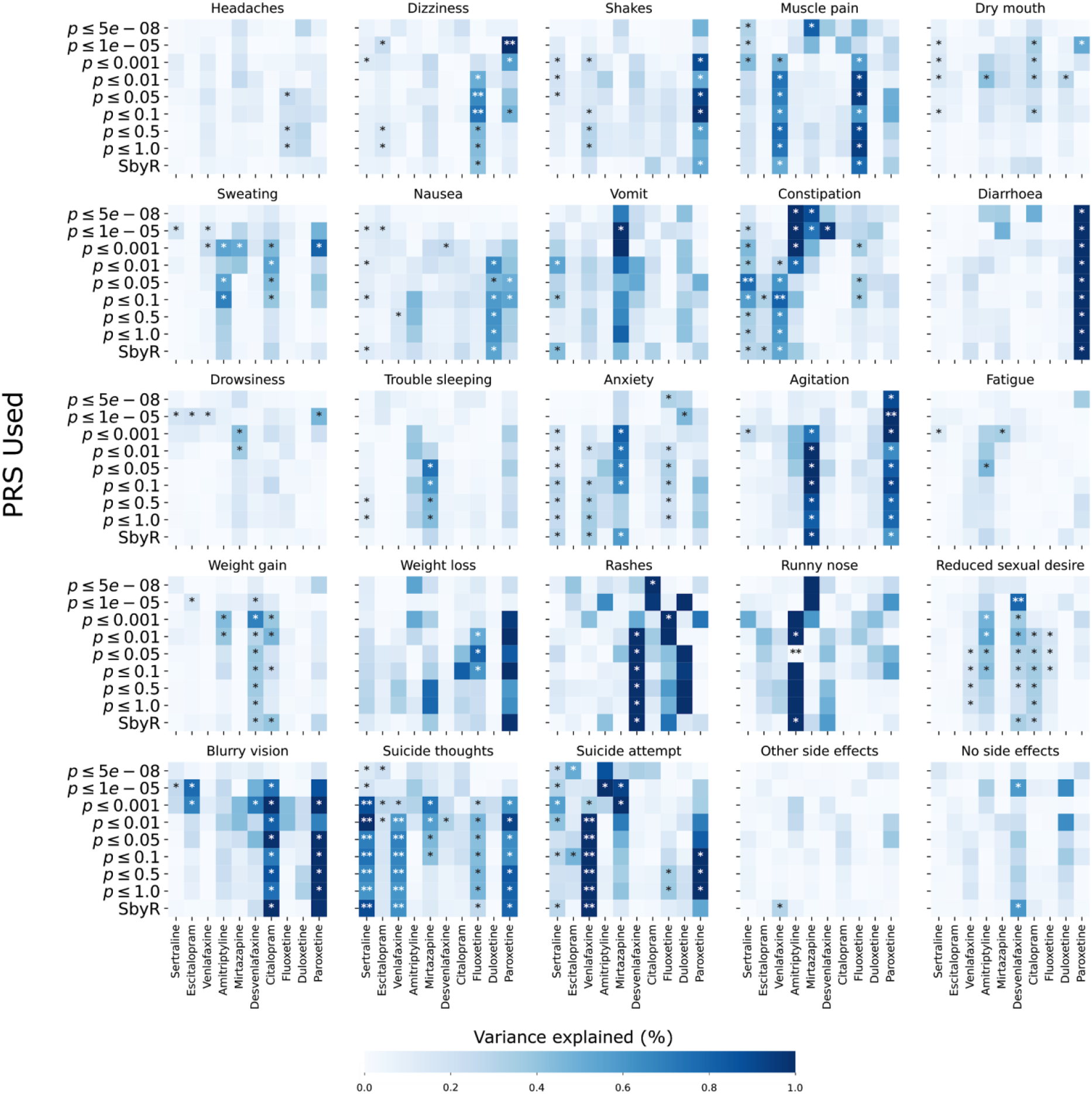
Depression PRS prediction of side effects. Heatmaps showing the results of depression PRS predicting side effects across antidepressant medication. Labels of the y axis show the p-value inclusion threshold for PRS calculation when using clumping and thresholding or *SbyR* when showing results for SBayesR PRS.

**Supplementary Figure 4.**
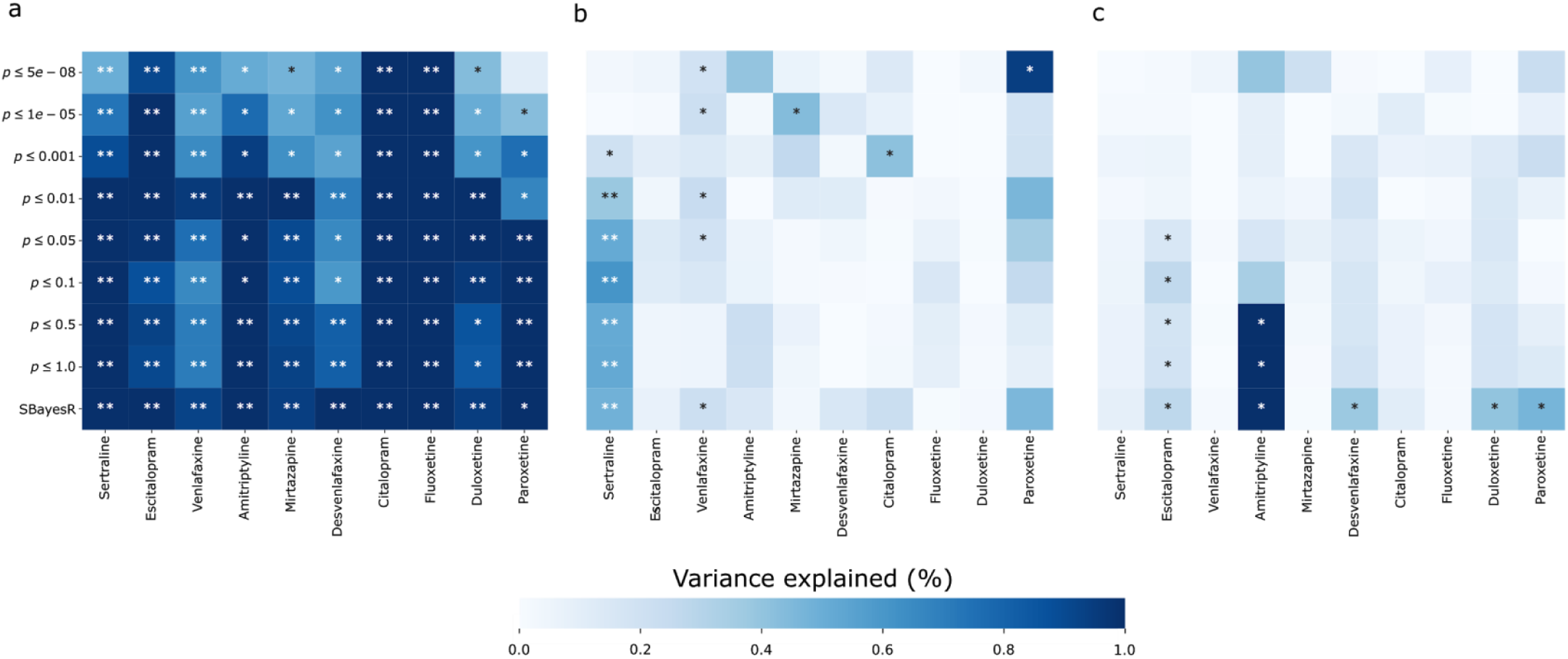
Genetic factors underlying headaches, insomnia and BMI. Heatmaps showing the results (variance explained) of BMI PRS predicting weight gain (a); headaches PRS predicting headaches (b) and insomnia PRS predicting insomnia (c) as side effects from antidepressants.

**Supplementary Figure 5.**
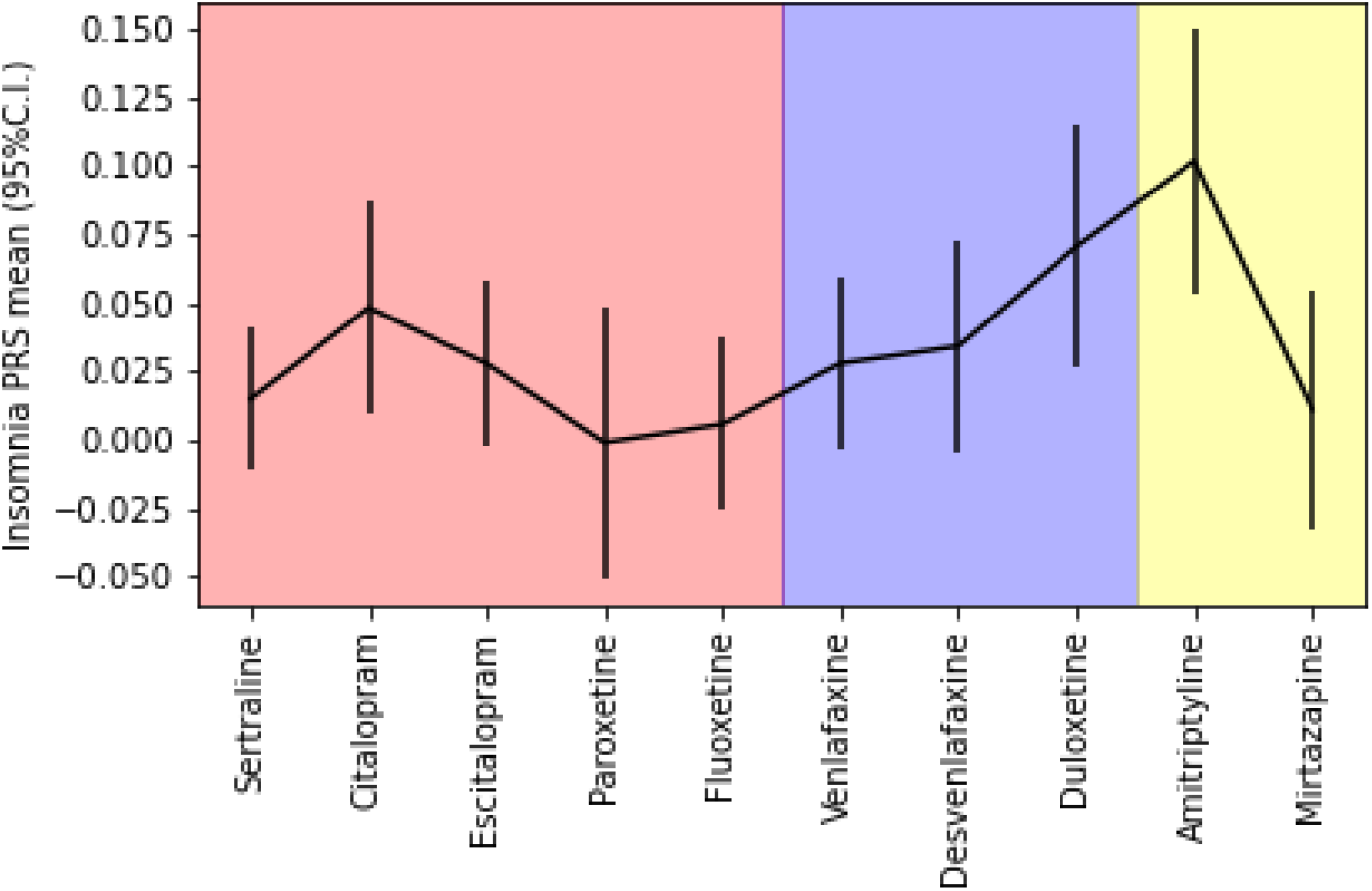
Insomnia PRS distribution across medications. Plot depicting the mean and 95% confidence interval for insomnia PRS across participants reporting taking the different medications under study. Note the higher insomnia PRS for participants taken amitriptyline; a pattern expected given its use to treat insomnia.

## Notes

### Competing Interest Statement

IBH has been commissioner of Australia National Mental Health Commission, Co-director of Health & Policy at the Brain & Mind Centre, University of Sydney. Led community-based and pharmaceutical industry-supported projects (Wyeth, Eli Lilly, Servier, Pfizer, AstraZeneca) focused on the identification and better management of anxiety and depression. Member of the Medical Advisory Panel for Medibank Private until October 2017. A board member of Psychosis Australia Trust. A member of the Veterans Mental Health Clinical Reference Group. Chief Scientific Advisor to and an equity shareholder in Innowell. AIC, AM, NGM, JGT, PAL, SEM, PL, JGT, NRW, EMB and MER have nothing to disclose.

### Author Declarations

The QIMR Berghofer Medical Research Institute Human Research Ethics Committee approved all questionnaires and research procedures (project 2118).

## References

1. Mathers C, World Health Organization (2008): The Global Burden of Disease: 2004 Update. World Health Organization.

2. Papakostas GI (2009): Serotonin Norepinephrine Reuptake Inhibitors: Spectrum of Efficacy in Major Depressive Disorder. Prim psychiatry 16.

3. Cipriani A, Furukawa TA, Salanti G, Chaimani A, Atkinson LZ, Ogawa Y, et al. (2018): Comparative Efficacy and Acceptability of 21 Antidepressant Drugs for the Acute Treatment of Adults With Major Depressive Disorder: A Systematic Review and Network Meta-Analysis. Focus 16: 420– 429.

4. Sansone RA, Sansone LA (2012): Antidepressant adherence: are patients taking their medications? Innov Clin Neurosci 9: 41–46.

5. Rosen RC, Lane RM, Menza M (1999): Effects of SSRIs on sexual function: a critical review. J Clin Psychopharmacol 19: 67–85.

6. Altman CA (2001, April): Effects of selective serotonin reuptake inhibitors on sexual function. Journal of Clinical Psychopharmacology, vol. 21. pp 241–242.

7. Lane RM (1997): A critical review of selective serotonin reuptake inhibitor-related sexual dysfunction; incidence, possible aetiology and implications for management. J Psychopharmacol 11: 72–82.

8. Bouwer CD, Harvey BH (1996): Phasic craving for carbohydrate observed with citalopram. Int Clin Psychopharmacol 11: 273–278.

9. Fava M (2000): Weight gain and antidepressants. J Clin Psychiatry 61 Suppl 11: 37–41.

10. Sussman N, Ginsberg DL, Bikoff J (2001): Effects of nefazodone on body weight: a pooled analysis of selective serotonin reuptake inhibitor-and imipramine-controlled trials. J Clin Psychiatry 62: 256–260.

11. Himmerich H, Minkwitz J, Kirkby KC (2015): Weight Gain and Metabolic Changes During Treatment with Antipsychotics and Antidepressants. Endocr Metab Immune Disord Drug Targets 15: 252– 260.

12. Thompson C (2002): Onset of action of antidepressants: results of different analyses. Hum Psychopharmacol 17 Suppl 1: S27–32.

13. Wichniak A, Wierzbicka A, Jernajczyk W (2012): Sleep and antidepressant treatment. Curr Pharm Des 18: 5802–5817.

14. Rush AJ, Armitage R, Gillin JC, Yonkers KA, Winokur A, Moldofsky H, et al. (1998): Comparative effects of nefazodone and fluoxetine on sleep in outpatients with major depressive disorder. Biol Psychiatry 44: 3–14.

15. Sharpley AL, Williamson DJ, Attenburrow ME, Pearson G, Sargent P, Cowen PJ (1996): The effects of paroxetine and nefazodone on sleep: a placebo controlled trial. Psychopharmacology 126: 50–54.

16. Brent DA (2016): Antidepressants and Suicidality. Psychiatr Clin North Am 39: 503–512.

17. Mihanović M, Restek-Petrović B, Bodor D, Molnar S, Oresković A, Presecki P (2010): Suicidality and side effects of antidepressants and antipsychotics. Psychiatr Danub 22: 79–84.

18. Read J, Williams J (2018): Adverse Effects of Antidepressants Reported by a Large International Cohort: Emotional Blunting, Suicidality, and Withdrawal Effects. Curr Drug Saf 13: 176–186.

19. Thakurta RG, Singh OP, Bhattacharya A, Mallick AK, Ray P, Sen S, Das R (2012): Nature of sexual dysfunctions in major depressive disorder and its impact on quality of life. Indian J Psychol Med 34: 365–370.

20. Baldwin DS (2001): Depression and sexual dysfunction. Br Med Bull 57: 81–99.

21. Wichniak A, Wierzbicka A, Walęcka M, Jernajczyk W (2017): Effects of Antidepressants on Sleep. Curr Psychiatry Rep 19: 63.

22. Goodwin GM (2006): Depression and associated physical diseases and symptoms. Dialogues Clin Neurosci 8: 259–265.

23. Kevin Hicks J, Sangkuhl K, Swen JJ, Ellingrod VL, Müller DJ, Shimoda K, et al. (2017): Clinical Pharmacogenetics Implementation Consortium Guideline (CPIC®) for CYP2D6 and CYP2C19 Genotypes and Dosing of Tricyclic Antidepressants: 2016 Update. Clin Pharmacol Ther 102: 37.

24. Tamminga WJ, Wemer J, Oosterhuis B, de Zeeuw RA, de Leij LF, Jonkman JH (2001): The prevalence of CYP2D6 and CYP2C19 genotypes in a population of healthy Dutch volunteers. Eur J Clin Pharmacol 57: 717–722.

25. Mrazek DA, Biernacka JM, O’Kane DJ, Black JL, Cunningham JM, Drews MS, et al. (2011): CYP2C19 variation and citalopram response. Pharmacogenet Genomics 21: 1–9.

26. Bråten LS, Haslemo T, Jukic MM, Ingelman-Sundberg M, Molden E, Kringen MK (2020): Impact of CYP2C19 genotype on sertraline exposure in 1200 Scandinavian patients. Neuropsychopharmacology 45. https://doi.org/10.1038/s41386-019-0554-x

27. Wigmore EM, Hafferty JD, Hall LS, Howard DM, Clarke T-K, Fabbri C, et al. (2020): Genome-wide association study of antidepressant treatment resistance in a population-based cohort using health service prescription data and meta-analysis with GENDEP. Pharmacogenomics J 20: 329– 341.

28. Li QS, Tian C, Hinds D, 23andMeResearchTeam (2020): Genome-wide association studies of antidepressant class response and treatment-resistant depression. Transl Psychiatry 10: 360.

29. Murray GK, Lin T, Austin J, McGrath JJ, Hickie IB, Wray NR (2021): Could Polygenic Risk Scores Be Useful in Psychiatry?: A Review. JAMA Psychiatry 78: 210–219.

30. Byrne EM, Kirk KM, Medland SE, McGrath JJ, Colodro-Conde L, Parker R, et al. (2020): Cohort profile: the Australian genetics of depression study. BMJ Open 10: e032580.

31. Howard DM, Adams MJ, Clarke T-K, Hafferty JD, Gibson J, Shirali M, et al. (2019): Genome-wide meta-analysis of depression identifies 102 independent variants and highlights the importance of the prefrontal brain regions. Nat Neurosci 22: 343–352.

32. Jansen PR, Watanabe K, Stringer S, Skene N, Bryois J, Hammerschlag AR, et al. (2019): Genome-wide analysis of insomnia in 1,331,010 individuals identifies new risk loci and functional pathways. Nat Genet 51: 394–403.

33. Lloyd-Jones LR, Zeng J, Sidorenko J, Yengo L, Moser G, Kemper KE, et al. (2019): Improved polygenic prediction by Bayesian multiple regression on summary statistics. Nat Commun 10: 5086.

34. Purcell S, Neale B, Todd-Brown K, Thomas L, Ferreira MAR, Bender D, et al. (2007): PLINK: a tool set for whole-genome association and population-based linkage analyses. Am J Hum Genet 81: 559–575.

35. Montejo-González AL, Llorca G, Izquierdo JA, Ledesma A, Bousoño M, Calcedo A, et al. (1997): SSRI-induced sexual dysfunction: fluoxetine, paroxetine, sertraline, and fluvoxamine in a prospective, multicenter, and descriptive clinical study of 344 patients. J Sex Marital Ther 23: 176–194.

36. Santarsieri D, Schwartz TL (2015): Antidepressant efficacy and side effect burden: a quick guide for clinicians. Drugs Context 4: 212290.

37. Jing E, Straw-Wilson K (2016): Sexual dysfunction in selective serotonin reuptake inhibitors (SSRIs) and potential solutions: A narrative literature review. Ment Health Clin 6: 191–196.

38. Stahl SM (1998): Mechanism of action of serotonin selective reuptake inhibitors. Serotonin receptors and pathways mediate therapeutic effects and side effects. J Affect Disord 51: 215– 235.

39. Safa M, Sadr S, Talischi F, Ghasem Boroujerdi F (2013): Study of effects of selective serotonin reuptake inhibitors on stages of sexual function in Iranian patients with major depressive disorder. Ther Adv Psychopharmacol 3: 306–313.

40. Frohlich PF, Meston CM (2000): Evidence that serotonin affects female sexual functioning via peripheral mechanisms. Physiol Behav 71: 383–393.

41. Stewart JW, McGrath PJ, Rabkin JG, Quitkin FM (1993): Atypical depression. A valid clinical entity? Psychiatr Clin North Am 16. Retrieved February 11, 2021, from https://www.ncbi.nlm.nih.gov/pubmed/8415233

42. Thaipisuttikul P, Ittasakul P, Waleeprakhon P, Wisajun P, Jullagate S (2014): Psychiatric comorbidities in patients with major depressive disorder. Neuropsychiatr Dis Treat 10: 2097– 2103.

43. Mimura M (2001): Comorbidity of depression and other diseases. Japan Med Assoc J 44: 225–229.

44. Coulehan JL, Schulberg HC, Block MR, Janosky JE, Arena VC (1990): Medical comorbidity of major depressive disorder in a primary medical practice. Arch Intern Med 150: 2363–2367.

45. Shelton CI (2004): Long-term management of major depressive disorder: are differences among antidepressant treatments meaningful? J Clin Psychiatry 65 Suppl 17: 29–33.

46. Caccia S (1998): Metabolism of the newer antidepressants. An overview of the pharmacological and pharmacokinetic implications. Clin Pharmacokinet 34. https://doi.org/10.2165/00003088-199834040-00002

47. Wharton S, Raiber L, Serodio KJ, Lee J, Christensen RA (2018): Medications that cause weight gain and alternatives in Canada: a narrative review. Diabetes Metab Syndr Obes 11: 427–438.

48. Planès S, Villier C, Mallaret M (2016): The nocebo effect of drugs. Pharmacol Res Perspect 4: e00208.

49. Wray NR, Ripke S, Mattheisen M, Trzaskowski M, Byrne EM, Abdellaoui A, et al. (2018): Genome-wide association analyses identify 44 risk variants and refine the genetic architecture of major depression. Nat Genet 50: 668–681.

50. Malhi GS, Bell E, Bassett D, Boyce P, Bryant R, Hazell P, et al. (2021): The 2020 Royal Australian and New Zealand College of Psychiatrists clinical practice guidelines for mood disorders. Aust N Z J Psychiatry 55: 7–117.

51. Cipriani A, Zhou X, Del Giovane C, Hetrick SE, Qin B, Whittington C, et al. (2016): Comparative efficacy and tolerability of antidepressants for major depressive disorder in children and adolescents: a network meta-analysis. Lancet 388: 881–890.

